# Impact on risk stratification of overlap syndrome phenotype (Brugada and Long QT type 3) in patients with E1784K mutation in SCN5A

**DOI:** 10.1101/2025.08.05.25333081

**Authors:** Fayçal Ben Abdallah, René Bun, Olivier Geoffroy, Pierre Roumegou, Frédérique Payet, François Wiart, Gaël Clerici, Josselin Duchateau, Maxime Churet

**Author notes:** **Funding:** None. **Data Availability Statement:** The data that support the findings of this study are available upon request from the participating institutions, in compliance with patient privacy regulations. **Corresponding author:** Dr Fayçal Ben Abdallah, Service de Cardiologie, CHU de La Réunion, Site Sud, Avenue François Mitterrand, 97448 Saint-Pierre, La Réunion, France. **Clinical Trial Disclosure:** NCT05274646. **Ethical Disclosure:** This study was approved by the Institutional Review Board of CHRU Saint Jacques in Besançon, France (ethics approval number: 2021-A03006-35).

## Abstract

**Introduction:** Risk stratification of patients with the SCN5A-E1784K mutation is challenging because they can express Brugada syndrome, long QT syndrome (LQTS), or both phenotypes (overlap syndrome). This study compared arrhythmic risk in patients with overlap syndrome versus single phenotype presentations.

**Methods:** We retrospectively enrolled patients aged 12 and older carrying the SCN5A-E1784K mutation. The primary outcome was major cardiac events (MCEs) defined as sustained ventricular arrhythmia, cardiopulmonary arrest and sudden cardiac death. Comparisons were made between patients expressing single phenotype (Brugada syndrome or LQTS type 3) and overlap syndrome.

**Results:** Forty-seven patients were enrolled (32 overlap group, 15 single phenotype group). MCE occurrence was 25% (8/32) in the overlap group and 6.7% (1/15) in the single phenotype group. Median follow-up was 156 months (single phenotype) and 127 months (overlap group). Despite a trend toward higher risk, comparison showed no statistically significant difference (OR 4.67, 95% CI: 0.53-41.4, p=0.275). Ajmaline testing reclassified 12% of patients, highlighting the importance of comprehensive diagnostic evaluation.

**Conclusion:** This study describes the largest cohort of SCN5A-E1784K mutation carriers reported. Although not statistically significant due to event rarity and sample size, results reveal a clear trend toward higher MCE prevalence in the overlap group. Ajmaline testing revealed underdiagnosis of overlap syndrome. Larger studies are needed for definitive conclusions.

## Introduction

Brugada syndrome (BrS) and long QT syndrome (LQTS) are inherited diseases known as cardiac channelopathies. Patients are mostly asymptomatics, with normal structural heart but with an increased risk of sudden cardiac death. The risk stratification in these patients is therefore both essential and challenging taken separately^1^ and this is particularly true when both conditions coexist in the same patient.

Indeed, among all types of LQTS, type 3 is notable for affecting the same gene as Brugada syndrome: the *SCN5A* gene. This gene encodes the principal cardiac Na+ channel isoform in the human heart, Nav1.5^2^ which is located on the cell membrane^3^. In LQTS, *SCN5A* mutations disrupt the fast Na+ channel inactivation, leading to a persistent inward Na+ current during the action potential plateau. This slows down the repolarization process, resulting in a prolonged QT interval on the surface electrocardiogram (ECG). Conversely, the ST elevation observed in Brugada syndrome may be linked to decreased initial opening of the Na+ channels in the right ventricular epicardial cells.

Therefore, a mixed phenotype can be observed with a single mutation in the *SCN5A* gene: LQTS type 3, Brugada syndrome, or both; a condition known as “overlap syndrome”^8–9^.

More specifically, the E1784K (p.Glu1784Lys) variant is the most prevalent mutation identified in patients expressing this syndrome^7^. This variant presents a particular challenge, as carriers of the E1784K mutation can exhibit features of both Brugada and LQTS simultaneously.

To date, the identification of a genetic mutation such as SCN5A-E1784K does not provide a clear prognostic element in a context where risk stratification is crucial, due to the rarity but severity of events such as sudden death. In fact, in patients known with a normal heart, Brugada syndrome could be related to up to 28% of sudden cardiac death cases^10^ and for resuscitated cardiac arrest, it could reach 5%-10%^11^ although these estimates are influenced by factors such as age and ethnic background. While LQTS1 and LQTS2 exhibit a higher frequency of cardiovascular events compared to LQTS3, events in LQTS3 are demonstrably more lethal. Nearly 20% of MCEs in LQTS3 are fatal, compared to a mortality rate below 5% in LQTS1 and LQTS2^12^.

Interestingly, this specific variant is significantly overrepresented among our patients, on Reunion Island, with an estimated prevalence of 9.26/100,000 inhabitants, in contrast to 2.26/100,000 inhabitants in the main french database (*cardiogen - région grand-ouest*).

Our local experience suggest a higher frequency of major cardiac events (MCEs) in patients expressing overlap syndrome phenotype (coexisting BrS and LQTS3) compared to those with a single phenotype (either BrS or LQTS3) although the scientific literature specifically comparing the occurrence of MCEs between these two groups is poor; thus demonstrating a real research challenge. In a prior study conducted at our center (not yet published), we compared MCE occurrences in patients with E1784K-mutated Brugada syndrome to those with Brugada syndrome caused by any other mutation in SCN5A gene. The E1784K variant itself did not appear to significantly worsen prognosis, reflecting the absence of impact of the genetic variant itself on risk stratification. However, it appeared that the expression of overlap syndrome phenotype may be associated with a higher risk.

The present study aims to compare MCEs occurrence (sustained ventricular arrhythmia, cardiopulmonary arrest and sudden cardiac death) in our cohort of patients with E1784K mutation in the *SCN5A* gene, between the group “single phenotype” (expressing either Brugada syndrome or LQTS type 3) and the group “overlap syndrome” (expressing both).

## Methods

### Study Design

The RISKOVER study is a hospital-based, monocentric, regional, cohort study, conducted at the University Hospital of Reunion Island.

### Patient selection

Patients eligible for this study were identified through the centralized E1784K variant screening program coordinated by the Genetics Department of the University of Nantes. This program encompasses a broad geographical area including the *Grand-Ouest* region and Reunion Island, both parts of *Cardiogen* network. Genetic testing for the SCN5A-E1784K mutation was performed on blood samples collected at the Nantes University Hospital^13^. This centralized approach ensured consistency and accuracy in patient identification. Patients followed within the Cardiology/Rhythmology Department of Reunion Island University Hospital were considered eligible for participation. Informed consent was obtained orally from all participants. For minors under the age of 18, parental consent was also secured to ensure ethical compliance. Exclusion criteria included age less than 12 years, inability to provide oral informed consent, and terminal illness. Annual follow-up visits were part of patient standard medical care^14^.

### Data collection

Data collection leveraged routine annual follow-up visits conducted in the Cardiology Unit of Reunion Island University Hospital. To identify suitable patients, we extracted Reunionese patients with the SCN5A-E1784K mutation. Medical records were reviewed to confirm the diagnosis of LQTS3 in the relevant patients, and to differentiate those with Brugada syndrome who had spontaneous type 1 from those identified through an ajmaline test. Subsequently, patients whose Brugada status had not been established (neither spontaneous type 1 nor pharmacological testing performed) underwent Ajmaline testing (1 mg/kg over 5-10 minutes)^15^, a diagnostic procedure using a sodium channel blocker to unmask latent Brugada syndrome, performed under a supervising rhythmologist with continuous ECG monitoring. Baseline corrected QT interval (QTc) was recorded at the time of diagnosis, along with medical history including any prior MCE. During routine follow-up visits, individualized rhythmology consultations addressed patient history, documented any previously unrecorded MCEs, and included a follow-up ECG with manual QT correction. Patients were followed from enrolment until April 2024, with regular reassessments integrated into their routine care.

### End points

The primary endpoint of this study was the occurrence of MCE, including sustained ventricular arrhythmia, cardiopulmonary arrest and sudden cardiac death. Syncopes were excluded because the etiologies can be varied and are often benign.

Secondary endpoints compared the incidence of MCE in subgroup analysis of patients with QTc ≥ 500 ms or spontaneous Brugada pattern.

### Sample size

We hypothesized that patients with “overlap syndrome” experience a significantly higher incidence of MCEs compared to those with a single phenotype. Specifically, we anticipated an incidence rate of 1 MCE per participant per year in the “single phenotype”^16^ group versus 2 MCEs per participant per year in the “overlap syndrome” group. This hypothesis was based on observed trends and preliminary data suggesting that the combined presentation of Brugada syndrome and LQTS type 3 phenotypes exacerbates the risk of severe cardiac events. To ensure study robustness and the valid findings, we accounted for potential challenges such as losing participants to follow-up. Assuming a 10% loss to follow-up, we determined a total sample size of 64 patients (32 per group) to achieve an 80% power and a 5% alpha-risk (Using the PASS® version 20 software). This provides a high level of confidence in detecting a statistically significant difference in the incidence of MCEs between the two groups.

### Statistical analysis

Baseline characteristics and clinical features were summarized using descriptive statistics. Comparisons between the overlap syndrome and single phenotype groups were performed using the tableone package in R. Categorical variables were analyzed using Fisher’s exact test due to small sample sizes, while continuous variables were compared using the Mann-Whitney U test given the non-normal distribution of the data. To estimate the association between group classification and the occurrence of MCEs, odds ratios (OR) with 95% confidence intervals (CI) were calculated using univariate logistic regression. No multivariate analysis was conducted due to the limited number of events. All statistical analyses were performed using R software version 4.0.0 under Windows. The analyses were conducted by the Centre for Clinical Investigation at Reunion Island University Hospital. No interim analyses were performed during the study to maintain the integrity of our results.

### Ethical approval

This study protocol was approved by an Ethics Committee, ensuring that all procedures adhered to the highest ethical standards. The authors confirm the accuracy and completeness of the data, as well as the fidelity of the study to the approved protocol. All adverse events were reported to the relevant health vigilance systems (e.g., care vigilance, pharmacovigilance, other regulatory frameworks) in accordance with current regulations. This comprehensive reporting mechanism is crucial for monitoring patient safety and managing any potential risks associated with the study.

## Results

Between January 2022 and April 2024, a total of 128 patients from the *Grand Ouest* database were screened. Among them, 79 reunionese patients with the SCN5A-E1784K mutation were eligible for inclusion. Ultimately, we included 47 patients, with the main reason for exclusion being the absence of ajmaline testing (n=27), which made it impossible to determine whether these patients had Brugada syndrome or not. Of the included patients, 32 were classified into the overlap group, and 15 into the single phenotype group (Figure 1). The composition of the single phenotype group was as follows: 10 patients had isolated Brugada syndrome, and 5 had isolated LQTS3. Baseline demographic and clinical characteristics are shown in Table 1. Among patients with Brugada syndrome, there were 6 with spontaneous type 1: one in the single phenotype group (10%) and 5 in the overlap group (15.6%). The median follow-up time was 156 months for the single phenotype group and 127 months for the overlap group.

**Figure 1.**
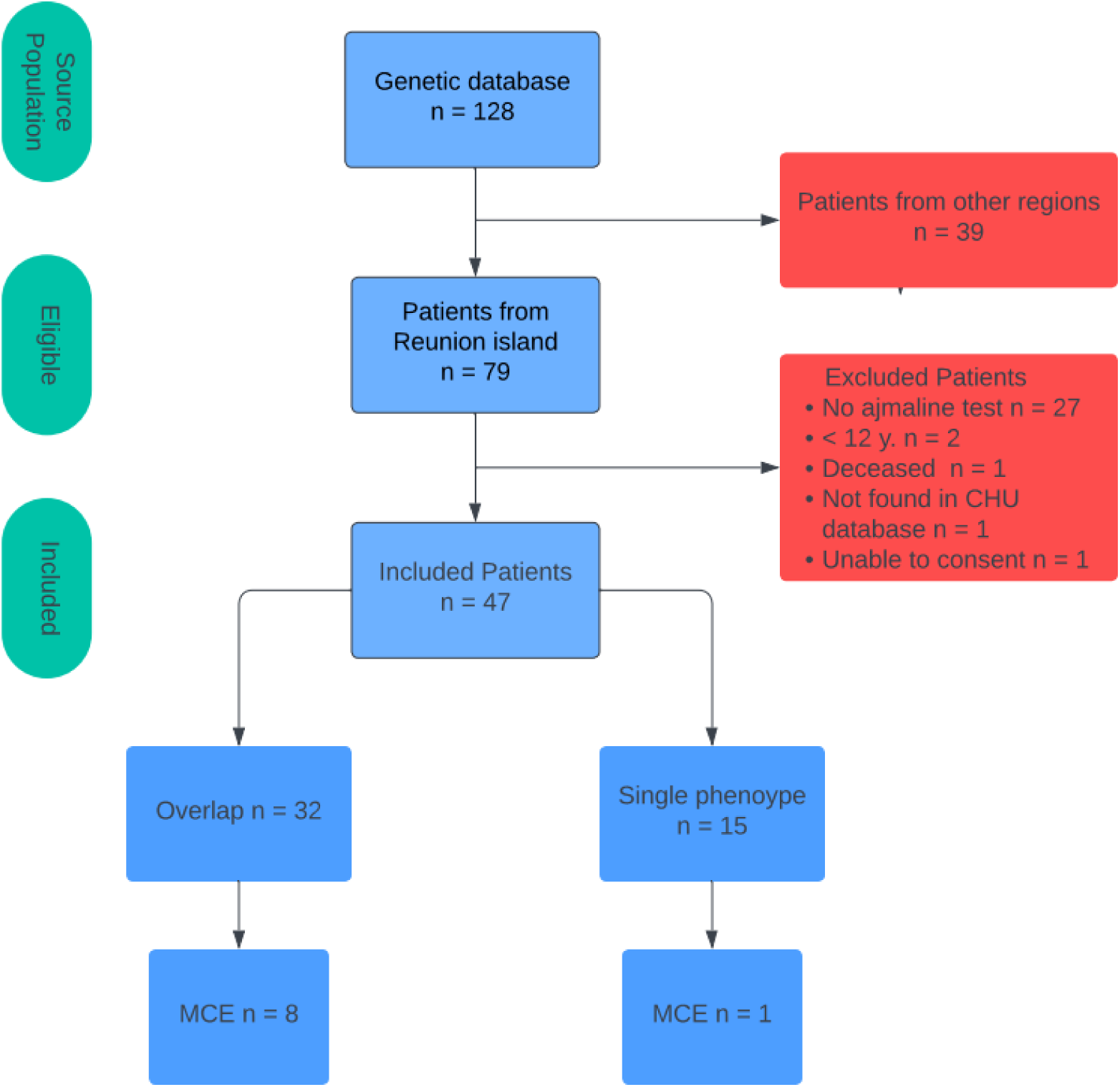
Flow Chart.

**Table 1.**
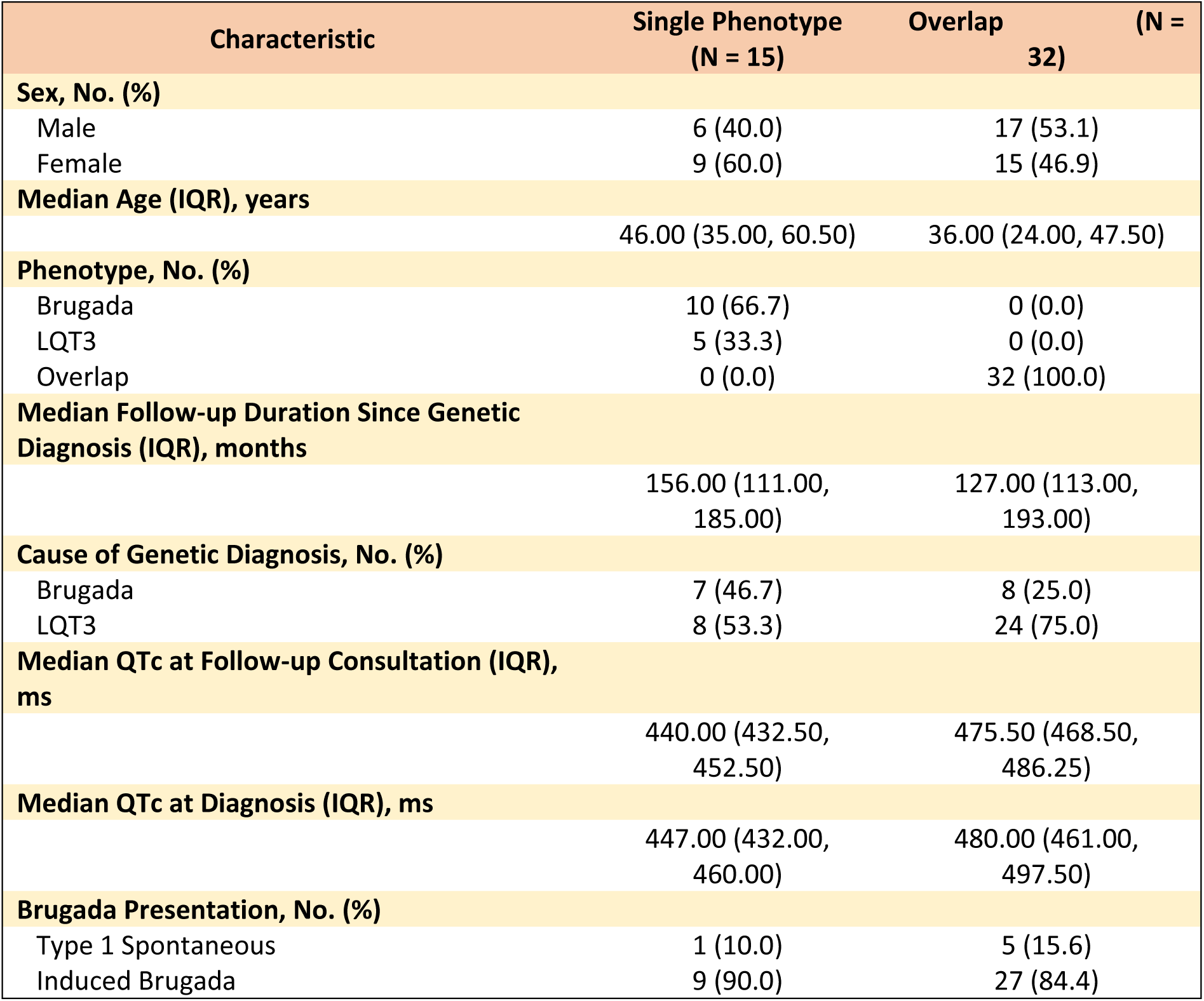
Baseline Characteristics of Patients.

**Table 2.**
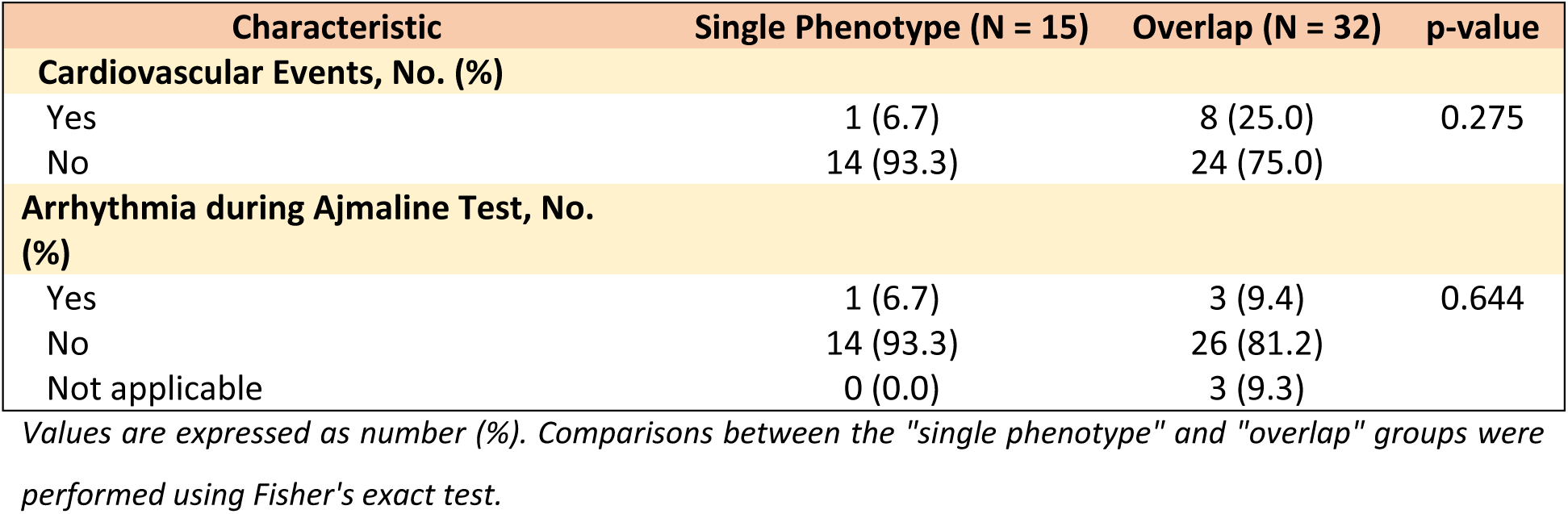
End Points.

| Characteristic | Single Phenotype (N = 15) | Overlap (N = 32) | p-value |
| --- | --- | --- | --- |
| Cardiovascular Events, No. (%) |  |  |  |
| Yes | 1 (6.7) | 8 (25.0) | 0.275 |
| No | 14 (93.3) | 24 (75.0) |  |
| Arrhythmia during Ajmaline Test, No. (%) |  |  |  |
| Yes | 1 (6.7) | 3 (9.4) | 0.644 |
| No | 14 (93.3) | 26 (81.2) |  |
| Not applicable | 0 (0.0) | 3 (9.3) |  |

Values are expressed as number (%). Comparisons between the "single phenotype" and "overlap" groups were performed using Fisher's exact test.

**Table 3.** Secondary End Point.

| Characteristic | QTc < 480 ms (N = 30) | QTc > 480 ms (N = 13) | p-value |
| --- | --- | --- | --- |
| Cardiovascular Events, No. (%) |  |  |  |
| Yes | 2 (6.7) | 2 (15.4) | 0.740 |
| No | 28 (93.3) | 11 (84.6) |  |
| Arrhythmia during Ajmaline Test, No. (%) |  |  |  |
| Yes | 3 (10.0) | 1 (7.7) | 0.644 |
| No | 24 (80.0) | 11 (84.6) |  |
| Not applicable | 3 (10.0) | 1 (7.7) |  |

Population characteristics stratified by qt time (threshold > 480 ms)

No statistically significant difference in the occurrence of MCEs between the overlap syndrome group and single phenotype group was observed. Specifically, 8 patients (25%) of the overlap syndrome group, experienced a major cardiovascular event, compared to 1 patient (6.7%) in the single phenotype group. The odds-ratio for this comparison was 4.67 (95% CI 0.53-41.4). However, the p-value for this comparison was 0.275, indicating no statistically significant difference between the groups.

In the secondary endpoints, we also compared the occurrence of MCEs between patients with QTc > 480 ms on the one hand and QTc < 480 ms on the other, to determine whether the possible excess risk might not be essentially mediated by a very long QTc interval. We thus identified 32 patients in the QTc < 480 ms group, 5 of whom suffered an MCE, and 15 patients in the QTc > 480 ms group, 4 of whom suffered an MCE. There was no significant difference in the occurrence of MCE between these two groups (p=0.57). In the QTc > 480 ms group, the 4 events were divided into 1 syncope and 3 ventricular fibrillations; in the QTc < 480 ms group, there were 2 ventricular tachycardias, 2 ventricular fibrillations and 1 syncope

However, none of the patients in the study experienced a fatal event during the follow-up period.

During the study, we observed several notable adverse events during the ajmaline testing procedure. Out of the 47 patients included in our cohort, 42 patients (89.4%) underwent pharmacological testing with ajmaline. The remaining five patients exhibited a spontaneous type 1 ECG pattern at baseline, eliminating the need for further pharmacological testing. Among the 42 patients who underwent ajmaline testing, we recorded four serious adverse events, representing an incidence rate of 8.5%. Specifically, two patients experienced ventricular fibrillation, another patient developed large monomorphic ventricular tachycardia, and the fourth patient encountered polymorphic ventricular arrhythmia.

## Discussion

We describe here the largest cohort of patients with the E1784K mutation of the *SCN5A* gene reported in the literature, in which we selected a stringent endpoint, providing a thorough assessment of the real risk associated with the SCN5A-E1784K mutation. We observed a trend towards higher prevalence of MCEs in the overlap group (8 events, 25%) compared to those with a single phenotype (1 event, 6.7%). This translates into an OR of 4.67, suggesting that individuals with overlap syndrome may be nearly five times more likely to experience MCEs. However, it is important to note that this difference is not statistically significant (p=0.275) likely due to the limited sample size and low event rates. Moreover, all of the events correspond to ventricular arrhythmias, which is reproducible with data from the literature^17^, and reinforces the confidence in the reliability of this trend.

Our study has several key strengths. Firstly, it provides valuable insights and contributes to the broader understanding of this rare mutation. The cohort established as part of the RISKOVER project on Reunion Island represents, from our knowledge, the largest cohort reported in the literature, while several international multicenter studies, including entire families investigating the SCN5A-E1784K mutation, have reported fewer mutated patients^18^. This highlights our contribution to the global research efforts on this rare genetic condition, yet dangerous. This large number of patients with the E1784K mutation underlines the specificity of the Reunionese population and the over-representation of this genetic variant on the island. An argument in favour of this high representation of the mutation could be a more marked search for Brugada syndrome on Reunion island than in other geographical areas, due to the island’s ethnic specificities^19^. But interestingly, the initial identification of LQTS type 3 led to genetic testing in the majority of cases (68%) in our cohort.

We tried to be as rigorous as possible in our judgment criteria, which is why we chose to exclude syncopal episodes, as we know their etiology is predominantly benign, despite their sometimes typical presentation during interrogation^20^. We also excluded ventricular arrhythmias that occurred during ajmaline tests because they were ‘induced.’ These choices can partly explain the non-significance of the results.

This study highlights the importance of close follow-up for patients carrying E1784K mutation, particularly those with overlap syndrome.

With regard to the occurrence of adverse events in the ajmaline test, in the literature, the estimated frequency of these events is around 0.5/100^21–23^. In our cohort, all four serious adverse events during ajmaline test occurred in the group overlap, with a ratio of 9.5/100. It is worth noting that the ajmaline tests were initially conducted over a duration of 5 minutes (previous recommendations suggested conducting the test for 5 to 10 minutes^24^). However, after deciding to conduct those tests over 10 minutes, as encouraged by the latest recommendations, no significant adverse events were observed^25^. Interestingly, four out of five overlap patients with spontaneous Brugada ECG type 1 experienced MCE, suggesting a potential sub-group with a higher risk^26^, requiring further investigations.

In addition, this study enabled us to identify 12 patients with previously undiagnosed overlap syndrome (because the ajmaline test had not been performed), suggesting underestimation of this association without routine ajmaline testing. Over 68.1% of patients were classified as overlap when the protocol was strictly followed, whereas only 42,5% initially. Given the qualitative analysis of our results, suggesting a higher risk of MCEs in overlap syndrome on one hand, and the risk of ventricular arrhythmia in ajmaline test one the other hand, a serious discussion should be opened about screening these mutated patients by pharmacological testing.

The question of treatment also arises for these patients with paradoxical indications, knowing the potential indication of treatment by antiarrhythmic class I for LQTS type 3 patients^27^ that would expose them at risk if they express Brugada syndrome. It remains unresolved, but recent study suggests benefit of mexiletine in SCN5A overlap syndrome.

The notable prevalence of severe ventricular arrhythmias within the overlap group emphasize the significant risk of sudden cardiac death associated with the combined phenotype. These findings underscore the importance of ongoing monitoring and research to elucidate the risks and clinical outcomes associated with different phenotypic expressions of this mutation. Further research and long-term follow-up are essential to optimize risk stratification and management strategies for patients with overlap syndrome, including medical treatment and defibrillator in primary prevention.

### Limitations

The first limitation is the lack of power. Although this cohort is historic, it remains too small to detect significant differences in major cardiovascular events between the overlap syndrome group and the single phenotype group. This limitation is compounded by the low event rates observed, which may obscure potential differences in risk. Additionally, the study’s monocentric design at a single hospital may limit the generalizability of the findings to other populations or settings. Furthermore, the observational nature of the study does not allow for causal inferences, and there may be unmeasured confounding factors influencing the results. Lastly, the follow-up duration, while substantial, may still be insufficient to capture the long-term risk of MCEs in these patients.

## Conclusion

This study described the largest cohort of SCN5A-E1784K mutation carriers to date, and identified a trend towards a higher risk of MCE in patients with overlap syndrome (Brugada and LQTS type 3) compared to those with isolated phenotypes (Brugada or LQTS type 3). Although the observed difference did not reach statistical significance due to the limited sample size, OR estimate revealed a marked over-representation of MCE, predominantly ventricular arrhythmias, in the overlap syndrome group. Additionally, ajmaline testing reclassified 12% of patients with overlap phenotype, emphasizing the importance of comprehensive diagnostic evaluation. These findings present serious clinical implications, because it arises questions about the indication of class I antiarrhythmic drugs in LQTS3 when co-expressing Brugada syndrome (overlap syndrome group) and the role of automatic implanted defibrillator in primary prevention in SCN5A-E1784K mutation carriers with overlap syndrome phenotype. Further research with a larger sample size is warranted to optimize knowledge and clinical management strategies for these patients.

## Data Availability

The data that support the findings of this study are available upon request from the participating institutions, in compliance with patient privacy regulations.

## Abreviations

BrS: brugada syndrome
CI: confidence interval
ECG: electrocardiogram
MCE: major cardiac event
OR: odds ratio
LQTS: long QT syndrome

